# A Pilot Study of Train-of-Four and Post-Tetanic Count Monitoring with the TetraGraph Electromyograph Compared to the TwitchView Monitor Electromyograph

**DOI:** 10.1101/2021.03.19.21249475

**Authors:** Logan Bussey, Srdjan Jelacic, Kei Togashi, Andrew Bowdle

## Abstract

**Background:** Quantitative twitch monitoring is recommended for avoiding residual neuromuscular blockade. Electromyograph twitch monitors are a form of quantitative twitch monitoring. The TwitchView electromyograph has been previously validated against “gold standard” mechanomyography, and may serve as a comparator for other monitors. We have previously shown that the GE electromyograph monitor overcounted twitches, frequently misinterpreting noise as a twitch. This is a pilot study to evaluate the performance of the TetraGraph electromyograph in comparison to the TwitchView electromyograph.

**Methods:** TwitchView and TetraGraph electrodes were applied to opposite arms of patients prior to induction of anesthesia. Post-tetanic count, train-of-four count and train-of-four ratio were then measured approximately every 5 minutes during an unrestricted general anesthetic. Measurements were not made for 10 minutes following neuromuscular blocking drugs or reversal agents.

**Results:** Eight patients were enrolled. The mean baseline train-of-four ratio was 1.02 (SD=0.04) for the TwitchView and 0.99 (SD=0.03) for the TetraGraph (p=0.22). Bland Altman analysis of all of the train-of-four ratio data found that average TwitchView train-of-four values were larger with a bias of 0.10. Train-of-four counts and train-of-four ratios were generally less when measured with TetraGraph than when measured with TwitchView.In 83% (209/253) of data pairs, the result from TetraGraph was less than the result from TwitchView and in 6% (16/253) of data pairs, the result from TetraGraph was greater than the result from TwitchView (p<0.0001). In 11% (28/253) of data pairs, the result from TetraGraph was the same as TwitchView [95%CI 7.35% 16.0%]. Evaluation of individual patient results confirmed the overall results. In some cases there were large discrepancies, such as 4 twitches reported by the TwitchView when the TetraGraph reported a post-tetanic count.

**Conclusions:** Users of the TetraGraph electromyograph should be aware that significant underestimation of post-tetanic-count, train-of-four count and train-of-four ratio may occur. This could result in administration of unnecessary reversal agents, excessive doses of reversal agents, or delay in extubation. We are undertaking a comparison of the TetraGraph monitor to mechanomyography to confirm the results of this pilot study.

## Introduction

Quantitative monitoring of twitch count (0-4 twitch responses evoked by a train-of-four stimulus) and train-of-four ratio (ratio between the fourth and first evoked twitch response) has been widely recommended for managing neuromuscular blockade and assessing the adequacy of recovery from blockade^1-3^. Mechanomyography is usually considered to be the laboratory “gold standard” for quantitative monitoring^3-5^, although it is not suitable for clinical monitoring and is not commercially available. Comparative studies have suggested that electromyography may be similar to mechanomyography or even “interchangeable” with mechanomyography^6,7^, however it does not necessarily follow that any specific electromyographic twitch monitor is in fact interchangeable with mechanomyography. Because electromyogram signals are of relatively small amplitude and electrical noise is ubiquitous, effective noise reduction is critical for the performance of an electromyogram-based monitor. If noise reduction is inadequate, noise may be incorrectly interpreted as electromyographic signal, or conversely, if the electromyogram signal is too aggressively filtered in an attempt to reduce noise, sensitivity for detecting the electromyogram signal may suffer. In a previous study of the GE electromyogram-based twitch monitor we found that electrical noise and artifacts were frequently misinterpreted as twitch^8^.

We have previously compared the TwitchView electromyograph (Blink Device Company, Seattle WA USA) to mechanomyography, and found that the TwitchView values were comparable to mechanomyography for train-of-four ratio^9^ and for train-of-four counting^10^. Based on these findings, we consider the TwitchView electromyograph to be a reference twitch monitor that can be used for comparison or validation of other twitch monitors. Only preliminary results have been reported for the TetraGraph electromyograph twitch monitor in comparison to the TOF Watch^11^ and Philips NMT acceleromyographs^12^. In both of these reports, the authors suggested that the TetraGraph appeared to have less sensitivity than the acceleromyographs under conditions of deeper levels of neuromuscular blockade, although most of the data presented concerned the train-of-four ratio rather than the post-tetanic count or the train-of-four count. The TetraGraph has also been used along with the TOF Watch acceleromyograph twitch monitor to measure the pain experienced during ulnar nerve stimulation in unmedicated volunteers^13^. There have been no published comparisons of the Tetragraph to other electromyographs or to mechanomyography. This study is a pilot study of the performance of the TetraGraph electromyograph. We compared TetraGraph post-tetanic count, train-of-four count and train-of-four ratio measurements to simultaneous measurements made with the TwitchView electromyograph.

## Methods

Our institutional review board approved this study and patients gave written, informed consent (UW HSD IRB approved 9/8/2017, University of Washington, Seattle WA 98195 USA). Patients with known neuromuscular abnormalities were excluded. The study was carried out between July 8, 2019 and August 6, 2019 at the University of Washington Medical Center. The twitch monitor data from this study were not used in the clinical care of the patients, and no health outcomes were measured.

We compared the TetraGraph to the TwitchView electromograph. The monitors were those available for purchase in Australia (TetraGraph) or the United States (TwitchView) at the time the study was conducted. TwitchView electromyography electrode arrays were used for the TwitchView monitor, and TetraGraph electrode arrays were used for the TetraGraph monitor; the electrode arrays for the monitors were placed on opposite arms. The amplitude of the train-of-four stimulus was set to 60 mA in all cases. No skin preparation was performed prior to attaching any of the electrodes. Temperature homeostasis was maintained in all patients through the use of active warming. End tidal CO2 was maintained between approximately 32 and 40 mmHg. The anesthesia technique including the choice of anesthetic and neuromuscular blocking agents was at the discretion of the anesthesia care team and included propofol, opioids (mainly fentanyl and hydromorphone), sevoflurane, isoflurane, rocuronium and vecuronium. Whenever possible, baseline measurements of train-of-four ratio were taken with each device after anesthetic induction but prior to initial administration of neuromuscular blocking drug. However, in some cases it was not possible to complete baseline measurements before the neuromuscular blocking drug was given. All train-of-four measurements were made in duplicate (i.e. two measurements were taken in a time span of less than 2 minutes) for each device approximately every five minutes from induction of anesthesia until just before emergence from anesthesia. Measurements were not made for 10 minutes following administration of neuromuscular blocking drugs or reversal agents in order to avoid periods when the extent of neuromuscular blockade was changing very rapidly.

### Endpoints of the study

The study endpoints were post-tetanic count, twitch count, and train-of-four ratio measured with TetraGraph and TwitchView monitors.

### Sample size calculations

As this was intended to be a pilot study to explore the performance of the TetraGraph and because of the scarcity of published data describing the performance of the TetraGraph monitor we did not attempt to calculate power. Because the tendency of the Tetragraph to undercount post-tetanic count and train-of-four count was clear and consistent, we did not find it necessary to recruit more than a small number of patients.

### Statistical analysis

Descriptive measures are presented as number (%) or mean ± SD and range, unless otherwise specified. The Bland-Altman method was used to plot the difference in train-of-four ratio for each twitch monitor against the mean of the two measurements. The limits of agreement are indicated as the interval of two standard deviations of the measurement differences on either side of the mean difference. A two-sample test of proportions was used to assess the twitch response between different monitors. Variables with a p-value less than 0.05 were considered statistically significant. All statistical comparisons were performed using STATA version 11.0 (StataCorp LP, College Station, Texas).

## Results

Eight patients were enrolled. Patient characteristics are shown in Table 1. All of the TwitchView and TetraGraph data pairs are plotted in Figure 1. Results for individual patients are shown in Figure 2.

**Table 1.**

|  |  |
| --- | --- |
| Number of subjects | 8 |
| Age (mean, SD, range) | 59.6, 10.6, 47-75 |
| Sex (F) | 6 (75%) |
| BMI (mean, SD, range) | 29.6, 6.2, 20.8-41.8 |
| ASA (1-5) |  |
| 1 | 0 |
| 2 | 1 |
| 3 | 7 |
| 4 | 0 |
| 5 | 0 |
| Duration of surgery (mean, SD, range) | 181, 96, 83-361 |
| Types of surgery |  |
| General | 4 |
| Gynecology | 4 |
| Individual ratio measurements per patient (with 4 twitches present across any compared devices) (mean, SD, range) | 16, 16, 2-51 |
| Individual count measurements per patient (1-3 twitch responses) (mean, SD, range) | 21.8, 14.5, 7-43 |
| Individual post-tetanic count per patient (mean, SD, range) | 9, 8, 2-28 |
| Number of patients receiving rocuronium | 2 |
| Number of patients receiving vecuronium | 6 |
| Number of patients receiving neostigmine | 3 |
| Number of patients receiving sugammadex | 5 |

**Figure 1.**
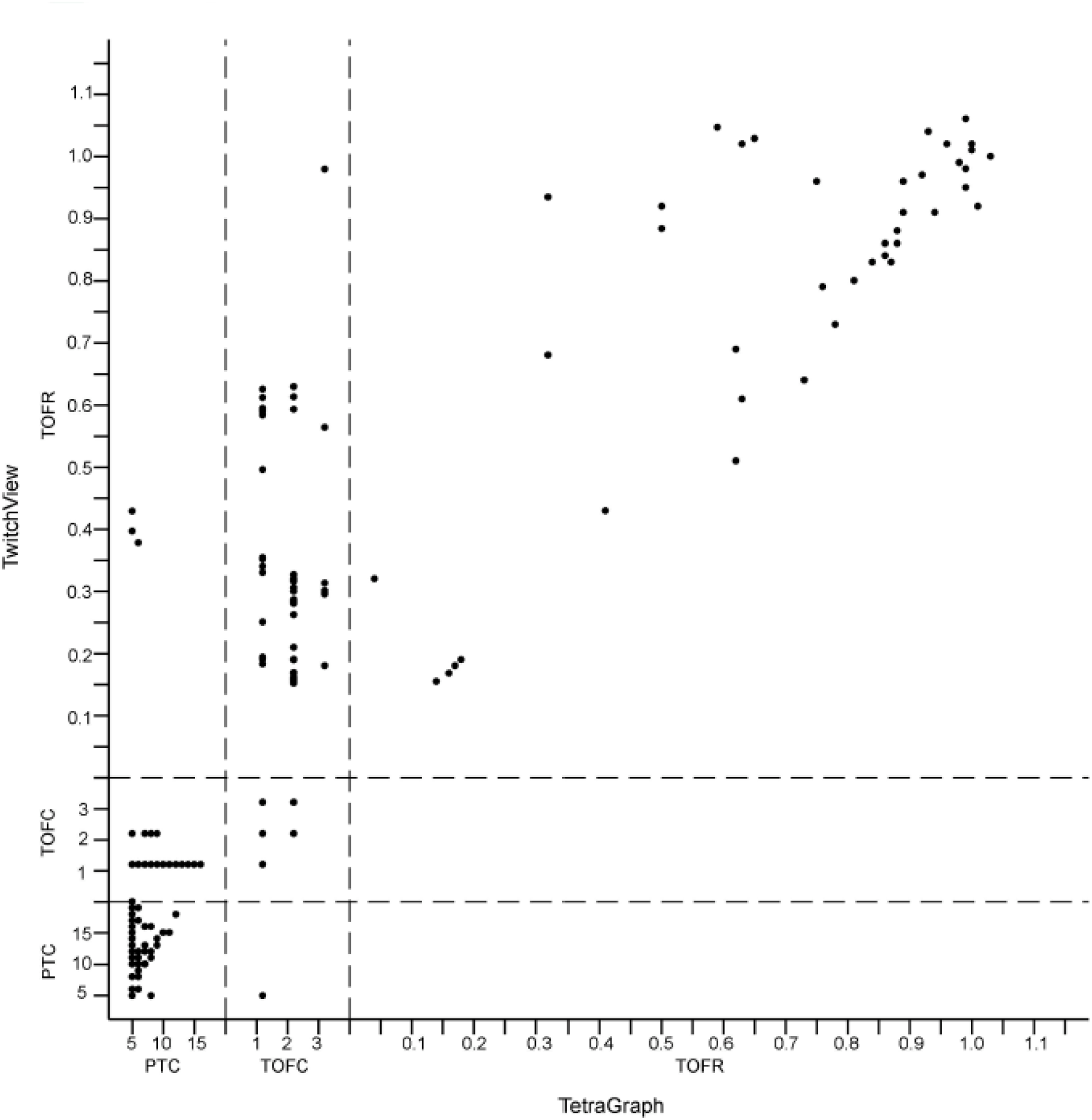
TetraGraph versus TwitchView. All 253 pairs of TetraGraph and TwitchView data points are shown. Because many pairs are identical, each dot may represent more than 1 pair. PTC=post-tetanic count. TOFC=train-of-four count. TOFR=train-of-four ratio.

**Figure 2.**
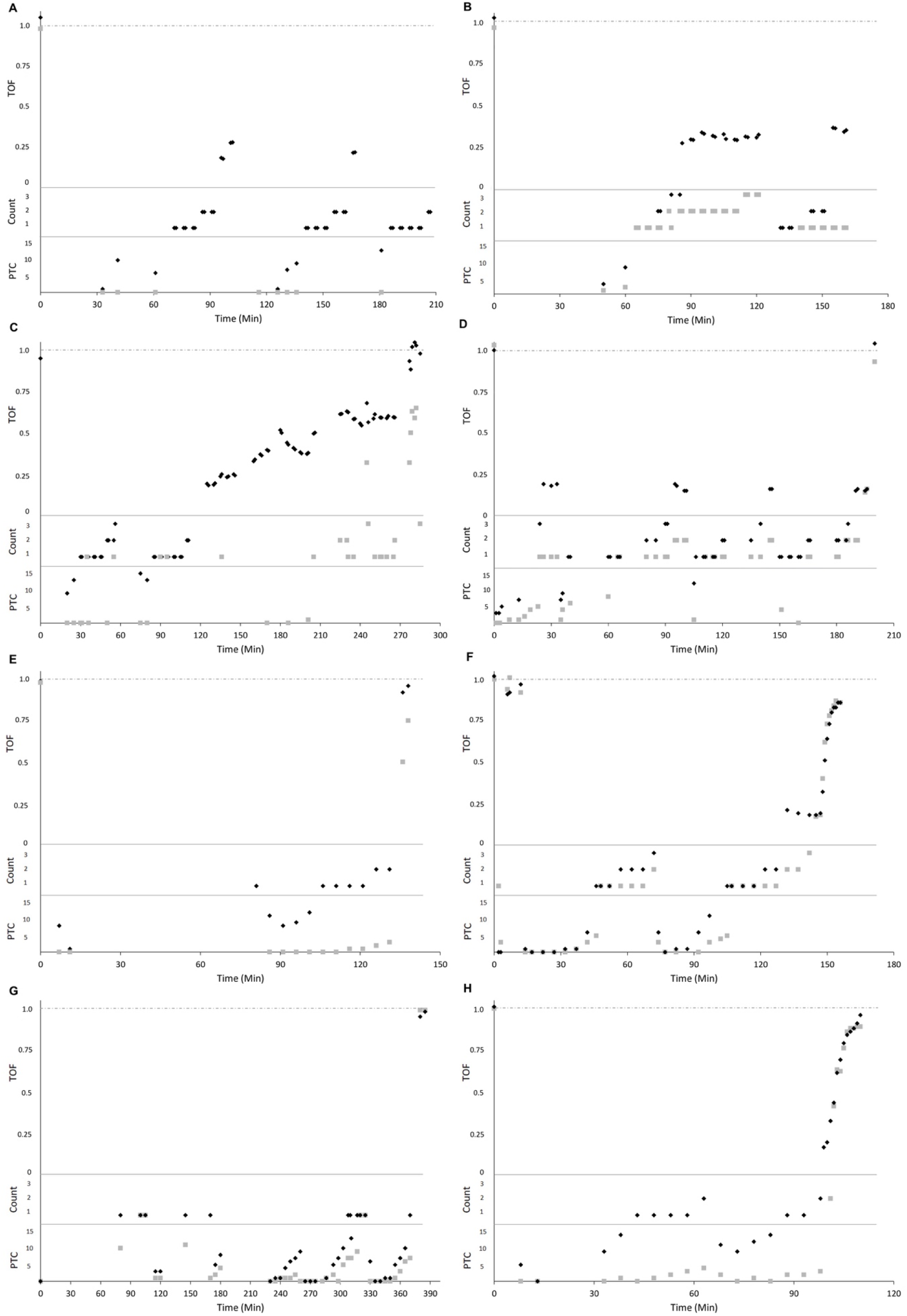
Individual Patient Data. Twitch responses for each patient are plotted against time. Dark diamonds are TwitchView data and light squares are TetraGraph data points. Baseline train-of-four ratio prior to administration of neuromuscular blocking drugs, when available, is plotted at time 0. There are a variety of explanations for “missing” data points. If a single point of a data pair is missing, one of the monitors was nonfunctional due to electrical interference (i.e. cautery) or some other technical issue. When both pairs of points are missing, either both monitors were nonfunctional, or more commonly some logistical issue prevented data collection (ie inability to access the monitors due to clinical activity in the operating room, positioning the patient for surgery, unavailability of the investigator to record data, etc). Also, data were not collected within 10 minutes of administration of neuromuscular blocking drugs or reversal agents. Otherwise, all data are reported as seen on the monitor screen, without any omissions or adjustments of results. See discussion section for further explanation.

In 6 of 8 patients, baseline train-of-four ratio was obtained prior to administering a neuromuscular blocking drug. The mean train-of-four ratio was 1.02 (SD= 0.04) for TwitchView and 0.99 (SD= 0.03) for TetraGraph (p= 0.22).

Following administration of a neuromuscular blocking drug, post-tetanic counts, train-of-four counts and train-of-four ratios were generally less when measured with TetraGraph than when measured with TwitchView. In 83% (209/253) of data pairs, the result from TetraGraph was less than the result from TwitchView and in 6% (16/253) of data pairs, the result from TetraGraph was greater than the result from TwitchView (p<0.0001). In 11% (28/253) of data pairs, the result from TetraGraph was the same as TwitchView [95%CI 7.35% 16.0%]. Bland-Altman analysis of pairs of train-of-four ratios (Figure 3) showed a bias of 0.10; limits of agreement −27.6,48.5), with train-of-four values being larger, on average, with TwitchView than with TetraGraph.

**Figure 3.**
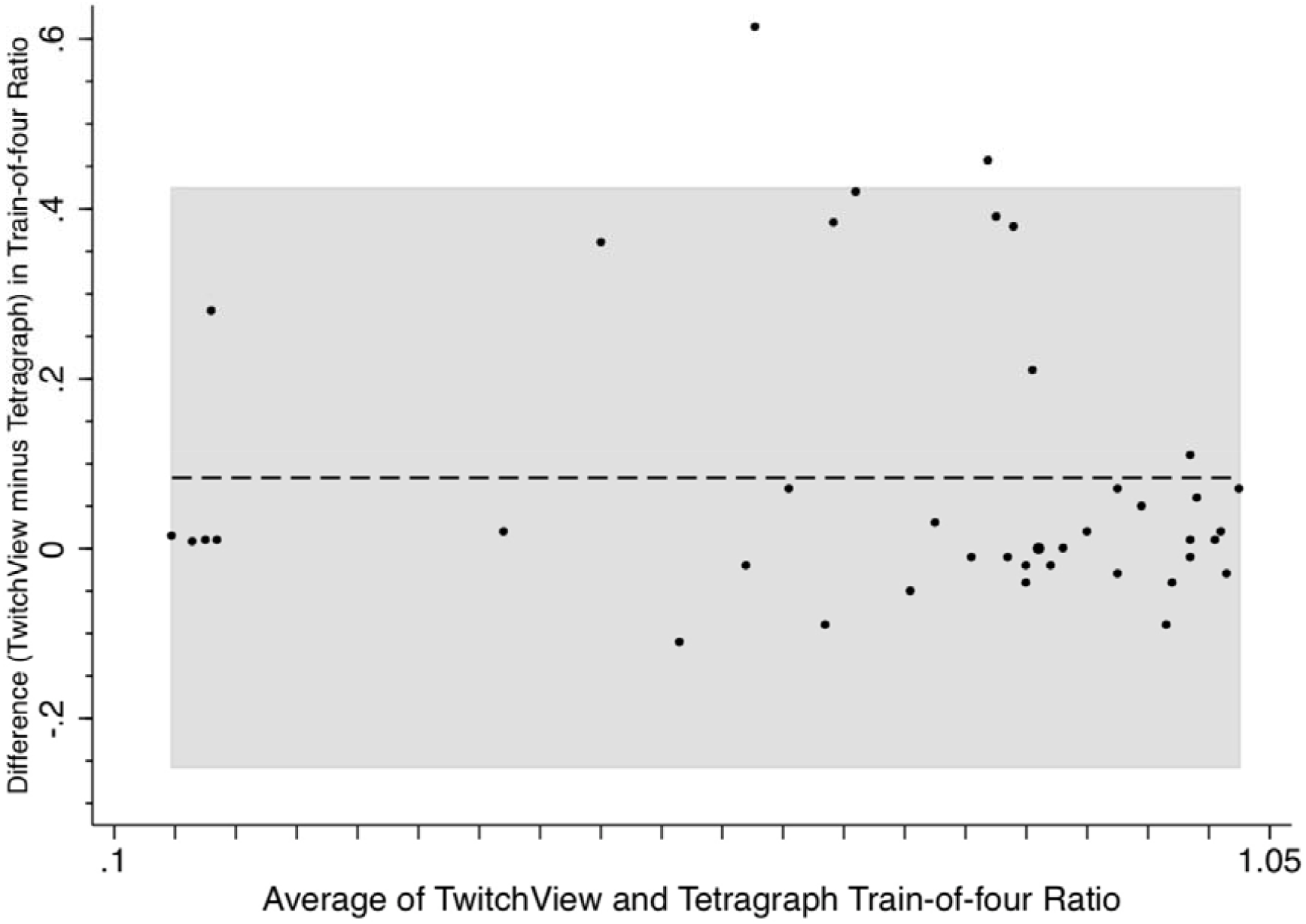
Bland Altman Plot of train-of-four ratio. The bias in the train-of-four ratio was 0.10, in the direction of larger train-of-four ratio for TwitchView compared to TetraGraph. The shaded area represents plus or minus 2 SD.

## Discussion

Prior to administration of a neuromuscular blocking drug, baseline twitch measurement with both TwitchView and TetraGraph obtained a train-of-four ratio close to 1.0, with no significant difference between the monitors. Following administration of a neuromuscular blocking drug, under a wide range of depth of neuromuscular blockade, the post-tetanic counts, train-of-four counts and train-of-four ratios were generally less with TetraGraph than with TwitchView. In many instances, the differences were substantial. This can be appreciated in Figure 1, in which all of the data pairs from the study are plotted. If the TwitchView and TetraGraph monitor had similar results, the data would be clustered along a diagonal line of identity. Instead, virtually all of the data points are above (to the left) of the line of identity, because TwitchView values for post-tetanic count, train-of-four count and train-of-four ratio are uniformly greater than TetraGraph. Inspection of the results for individual patients confirms this pattern. Panel C in Figure 2 is illustrative. From about 90 minutes until just before the end of data collection, the TwitchView reported 4 twitches with a gradually increasing train-of-four ratio, while the TetraGraph reported either a post-tetanic count or a train-of-four count less than 4. Following reversal, the TwitchView reported a train-of-four ratio of approximately 1.0, while the highest train-of-four ratio reported by the TetraGraph was <0.75. Individual patient results shown in panels A, B, D, E, G and H also demonstrate that TetraGraph twitch response values were generally less than the TwitchView twitch response values. Inspection of the data reveals that in many instances, the TetraGraph reported a post-tetanic count of zero at a time when the TwitchView reported one or more twitches. Panel F represents an exception to this pattern, in which there was a relatively closer correspondence between the monitors, although when the monitors differ, the TetraGraph values are again less than the TwitchView values.

Considering that TwitchView was previously shown to give results similar to mechanomyography ^9,10^, and therefore represents a reasonable reference for comparison to other monitors, it appears that the TetraGraph frequently underestimated the post-tetanic count, the train-of-four count and the train-of-four ratio. Our results are generally compatible with preliminary reports of the TetraGraph monitor in comparison to TOF Watch^11^ or Philips NMT^12^ acceleromyographs, in which the authors reported that the TetraGraph monitor appeared to be less sensitive under conditions of deeper levels of neuromuscular blockade; however the data presented in these preliminary reports were limited.

A technology challenge unique to electromyography based quantitative twitch monitors is measuring a relatively small electrical signal (several millivolts) in the presence of large electromagnetic interference. The TwitchView employs an EMG circuit with three electrodes. Two electrodes are used to detect the EMG signal while the third electrode actively cancels electrical noise (this technique maximizes the signal-to-noise ratio with minimal distortion of the EMG signal)^14^. The Tetragraph monitor uses two electrodes in its EMG circuit, and consequently has to rely on filtering to remove electrical noise^11^, with the possibility that significant portions of the desired signal are removed. A practical consequence of this approach may be reduced sensitivity for detecting small twitches, as we found in the present study, and has been previously reported by Nemes et al^11^. While Nemes et al conclude that the sensing threshold can be adjusted to improve detection of small twitches, this adjustment will not fix the underlying problem of poor signal-to-noise due to filtering. It is possible that simply adjusting the sensing threshold without improving the signal-to-noise will cause the system to mistakenly identify electrical noise as a twitch, a problem we and others have observed with the GE system electromyograph based twitch monitor^8^.

An assumption is sometimes made that all electromyography-based twitch monitors should obtain similar results because their underlying technology is the same. Our studies have shown that this assumption is not correct. We showed previously that the GE electromyography based twitch monitor did not effectively distinguish signal from noise, resulting in spurious twitch counting, for example counting four twitches when none were present^8^. In the current study we have shown that the TetraGraph twitch monitor has the opposite problem from the GE twitch monitor. The TetraGraph monitor frequently fails to detect twitches probably because of the methods utilized for removing electrical noise.

There are a number of limitations to our study. The number of subjects studied was small. However, because the differences between the TetraGraph and the TwitchView monitors were so pronounced, and this study was intended to be a pilot study, we terminated the study when it became apparent that the TetraGraph monitor was frequently undercounting twitches. We deliberately did not standardise the anesthetic care or neuromuscular blocker administration, in order to obtain results that would resemble routine anesthetic care. One of the disadvantages of this approach was that in some cases, patients were managed under deep block for most of the surgical procedure, limiting the opportunity to measure train-of-four responses. Because of this, some patients contributed more train-of-four data points than others. We did not prepare the skin prior to applying electrodes, because in our experience this measure is not widely used in anesthesia practice. We previously compared electromyograph signal quality on healthy volunteers before and after skin preparation using either alcohol or mild skin abrasion (unpublished results). Skin impedance was reduced with skin preparation, but there was not an appreciably larger amplitude signal. There can be arm-to-arm differences in twitch response, probably due to effects of the blood pressure cuff (on a single arm) on distribution of neuromuscular blocking drugs, however these differences seem most pronounced immediately after drug administration, and are less noticeable over a longer period of time. In a previous study, we found that overall results were not affected by measuring twitch from both arms^9^, confirming the findings of other investigators^15^.

A possible criticism of our study is that we did not use mechanomyography as a comparison for both TwitchView and TetraGraph monitors. Because we have previously validated the TwtichView train-of-four ratio^9^ and train-of-four counting^10^ performance in comparison to mechanomyography, we would argue that the TwitchView is a reasonable reference for comparison to other twitch monitors. Despite the shortcomings associated with acceleromyography^16^, the TOF Watch acceleromyography based twitch monitor has been used as the reference twitch monitor in many studies of twitch monitors^17^. We would argue that the TwitchView electromyography-based monitor, having been validated against mechanomyography, is a superior reference twitch monitor because it does not produce baseline train-of-four ratio greatly in excess of 1.0, as is commonly seen with acceleromyography. However, we are undertaking a study to compare the TetraGraph to mechanomyography as a further confirmation of our findings.

The clinical implications of underestimating and overestimating twitch are distinctly different. When twitch is overestimated (as with the GE electromyography-based twitch monitor), clinicians may be led to administer more neuromuscular blocking drug when none is needed, or to judge that patients are recovered when in fact they are not. This may result in the complications that may occur with residual neuromuscular blockade. When twitch is underestimated (as with the TetraGraph electromyography-based twitch monitor), clinicians may be led to administer less neuromuscular blocking drug than necessary to produce a particular depth of blockade, however it is unlikely that a patient will be regarded as having recovered when they have not. Underestimation of twitch may result in administration of unnecessary reversal agents, larger doses of reversal agents than necessary, or delay in extubation.

In conclusion, we found that the TetraGraph monitor was similar to the TwitchView monitor in measuring a normal train-of-four ratio under baseline conditions, prior to administration of neuromuscular blocking drugs, but significantly underestimated twitch response over a wide range of depth of neuromuscular blockade.

## Data Availability

Raw data are available from the authors on request.

